# Prevalence and determinants of problematic khat use in Ethiopia

**DOI:** 10.64898/2026.10.26.26364894

**Authors:** Awoke Mihretu, Wubalem Fekadu, Eyerusalem Solomon, Azeb Asaminew Alemu, Kassahun Habtamu, Solomon Teferra, Abebaw Fekadu

## Abstract

**Background:** Evidence on khat use prevalence is substantial in Ethiopia and elsewhere. However, problematic khat use remains largely unstudied because the concept has not been clearly defined or consistently measured. This study estimated the prevalence and determinants of problematic khat use among Ethiopian adults.

**Methods:** This descriptive cross-sectional analysis used baseline data from our Prospective khat cohort study conducted in Butajira and adjacent districts in Central Ethiopia Regional State. We included 504 khat-user participants who had been chewing khat within the previous five years. We assessed problematic khat use using the locally developed and validated 17-item Problematic Khat Use Screening Tool (PKUST-17). A score of ≥17 indicated problematic use. Binary and multivariable logistic regression models identified associated factors.

**Results:** Among the 504 khat-user participants, most were male (82.3%), married (85.0%), and rural residents (70.8%). The prevalence of problematic khat use was 19.6%. Daily or near-daily khat users had eightfold higher odds of problematic use than occasional users (AOR = 8.05, 95% CI: 3.75–17.24). Stressful life events were associated with a 3.48-fold increase (AOR = 3.48, 95% CI: 1.56–7.76), and depressive symptoms with a 3.70-fold increase in odds (AOR = 3.70, 95% CI: 2.00–6.75).

**Conclusion:** Problematic khat use affects approximately one in five khat users in Ethiopia. It is strongly associated with frequent khat use, stressful life events, and depression. Longitudinal studies are needed to clarify causal pathways and evaluate culturally appropriate prevention and treatment interventions.

## Background

Khat (*Catha edulis*) is a psychoactive plant with stimulant properties mainly attributed to cathinone and cathine (Kalix, 1990). Khat use is deeply embedded in social and cultural practices, particularly in Ethiopia, Yemen, Somalia, Kenya, and among migrant communities originating from these regions.

High levels of khat use have been reported in producing countries, including Kenya (31.5%), Yemen (67.9%), and Somalia (59%) ( Elmi, 1983; Numan, 2004; Ongeri et al., 2019). Khat use has also been reported among migrant populations, including Somali communities in the United Kingdom, where approximately 33% of individuals reported current use (Patel et al., 2005).

Ethiopia is one of the world’s major producers and consumers of khat. Nationally representative surveys indicate that khat use remains common among Ethiopian adults, increasing from approximately 16% in earlier national estimates to about 23% in recent surveys (Ethiopian Public Health Institute (EPHI), 2025; Teklie et al., 2018).

Most existing studies have focused on describing khat use rather than examining the transition from use to problematic khat use and its associated harms. Problematic khat manuscriptuse has increasingly been recognised as a clinically relevant condition characterized by psychological dependence, impaired control, withdrawal-like symptoms, and functional impairment. Historically, the possibility of khat dependence was debated due to limited evidence regarding physiological dependence and withdrawal. However, observational and cross-sectional studies have consistently reported symptoms following reduction or cessation of khat use, including craving, irritability, mild depressive symptoms, sleep disturbances, reduced energy, and continued use to relieve unpleasant experiences (Al-Motarreb et al., 2002; Jemal et al., 2015; Kalix & Braenden, 1985; Mihretu et al., 2017). Although khat withdrawal symptoms are generally less severe than those associated with substances such as alcohol or opioids, problematic use may result in meaningful psychological, social, and occupational consequences.

Available evidence suggests substantial variability in the prevalence of problematic khat use across populations, partly due to differences in definitions and measurement approaches. In Ethiopia, reported prevalence estimates range from approximately 20% in community-based samples to more than 70% among selected groups of frequent users. Similarly, studies from Yemen, Saudi Arabia, and migrant populations have reported psychological dependence among approximately 30–50% of khat users depending on the population and assessment method used(El-Setouhy et al., 2016a; Griffiths et al., 1997; Kassim et al., 2013). These findings suggest that a proportion of khat users develop problematic patterns associated with dependence symptoms and functional impairment.

Despite growing recognition of problematic khat use, its epidemiology remains difficult to establish due to the lack of culturally appropriate measurement tools. Previous studies have used diverse approaches, including the Severity of Dependence Scale (SDS), DSM-IV/DSM-5 substance use disorder criteria, and Drug Abuse Screening Test (DAST-10). However, many of these instruments were originally developed for other substances and may not adequately capture the behavioural, social, and cultural dimensions specific to problematic khat use. In Ethiopia, existing tools for assessing harmful khat use or khat use disorder have been developed mainly among student populations and often relied on adaptation of existing substance-use measures rather than comprehensive exploration of khat-specific experiences (Duresso et al., 2016). Consequently, differences in measurement approaches have contributed to inconsistent prevalence estimates and limited utility to inform policy.

To address this gap, our research team developed and validated the Problematic Khat Use Screening Tool (PKUST-17), a culturally relevant instrument designed to assess problematic khat use in Ethiopia(Mihretu et al., 2019, 2020, 2022b). However, despite advances in measurement, evidence regarding the prevalence and determinants of problematic khat use in community-based populations remains limited. Understanding problematic khat use at the community level is essential because harmful patterns of consumption may contribute to mental health problems, reduced quality of life, and social and occupational impairment. Therefore, this study aims to investigate the prevalence of problematic khat use and identify determinants among adults in Ethiopia.

## Methods

### Study design and setting

This study uses baseline data from our Butajira Khat Cohort Study, a community-based prospective cohort conducted in Butajira and adjacent rural districts in the Central Ethiopia Regional State. The overall cohort enrolled 923 adults to investigate the physical, mental, and socioeconomic consequences of khat use. The parent cohort methods have been reported elsewhere(Mihretu et al., 2026).

### Study population

The source population comprised adults residing in the Butajira Health and Demographic Surveillance System (HDSS) area. The Butajira HDSS is a population-based longitudinal cohort in Ethiopia that tracks demographic and health information over time. The surveillance system covers approximately 54,000 residents and is governed by the School of Public Health at Addis Ababa University. For our multidimensional harms of khat use study, we enrolled 923 randomly selected participants. Participants were eligible if they were aged 18 years or older, permanent residents of the HDSS area, and provided with informed consent.

### Sampling and sample size

The sample size for the parent cohort was calculated to examine physical, mental health, and economic outcomes associated with khat use. Assuming that 50% of adults would be regular khat users, based on previous studies from the area, the required sample was estimated using a single population proportion formula with a 95% confidence level, 80% power, a 1:1 exposed-to-unexposed ratio, and 15% allowance for loss to follow-up, yielding a minimum sample of approximately 400 participants. We randomly selected eligible participants from the Butajira HDSS population register. The current study included all 504 khat users available at baseline.

### Data collection instruments

Problematic khat use: We assessed the primary outcome using the Problematic Khat Use Screening Test (PKUST-17), a locally developed and validated 17-item screening instrument. The tool was developed through systematic literature review, item generation, cognitive interviewing, pilot testing, and psychometric validation. PKUST-17 has demonstrated good reliability and content, semantic, and construct validity, with a unidimensional factor structure. Item response theory analyses showed optimal performance for detecting moderate levels of problematic khat use. A total score of 17 or above was used to classify participants as having problematic khat use (Mihretu et al., 2022a, 2022b, 2025). Khat use frequency: Khat chewing frequency was categorized as daily or almost daily use (≥4 days per week) and occasional use (1–3 days per month).

Depressive symptoms: Depression was measured using the Patient Health Questionnaire-9 (PHQ-9). The PHQ-9 has been widely validated in Ethiopia and demonstrates acceptable psychometric properties. A score of 10 or higher indicated clinically significant depressive symptoms(Gelaye et al., 2013).

Sociodemographic characteristics: Information on age, sex, marital status, educational attainment, employment status, and place of residence was collected using a structured questionnaire. Household wealth index was constructed from household asset ownership using principal component analysis and categorised into wealth quintiles (Mihretu et al., 2026).

Stressful life events: Exposure to stressful life events was measured using a 12-item checklist covering events such as bereavement, divorce, theft, legal disputes, and major financial problems during the previous 30 days (Brugha & Cragg, 1990). Responses were dichotomised into no stressful life event and one or more stressful life events.

Household food insecurity: Household food insecurity was assessed using the Household Food Insecurity Access Scale (HFIAS), a standardized nine-item questionnaire developed by Coates et al. (2007) (Coates et al., 2007). The HFIAS measures anxiety about food supply, insufficient food quality, and inadequate food intake over the previous four weeks. For this analysis, food security status was dichotomized: food secured included households that were food secure or mildly food insecure, while food insecure included households that were moderately or severely food insecure.

Chronic physical illness: Participants were asked whether a healthcare professional had ever diagnosed them with hypertension, diabetes mellitus, or heart disease. A positive response to any condition was classified as having a chronic physical illness.

Other substance use: Current alcohol and tobacco use were assessed based on self-reported use during the previous 30 days.

## Data collection procedure

Data were collected through face-to-face interviews using standardised questionnaires in participants’ homes, conducted by experienced Butajirra HDSS staff trained for the study. Supervisors monitored data quality through daily field supervision, completeness checks, and consistency verification before data entry.

## Data analysis

Descriptive statistics summarised participants’ characteristics. The primary outcome was problematic khat use, categorised as PKUST-17 positive or negative. We used binary logistic regression to identify factors associated with problematic khat use. We entered variables with potential associations into the multivariable model to estimate adjusted odds ratios (AORs) with 95% confidence intervals. We declared statistical significance at p < 0.05. All statistical analyses were performed using Stata version 19.0.

## Ethical considerations

The study was approved by the Institutional Review Board of the College of Health Sciences, Addis Ababa University (Reference: 023/24/Psy). The researchers obtained written informed consent from all participants before enrollment. The study followed national and international ethical guidelines for research involving human participants.

## Results

Most participants were male (82.3%), married (85.0%), and rural residents (70.8%). Among the 504 participants who chewed khat, 99 (19.6%) had problematic khat use (PKUST-17 score >17). Nearly half reported daily or almost daily khat use (48.8%). Most participants had secondary education (44.7%), were farmers (50.4%), and lived in food-insecure households (80.2%). Problematic khat use was notably more common among men (22.9%), participants in the richest wealth quintile (28.2%), daily/almost daily khat users (39.7%) compared with occasional users (5.6%), current tobacco smokers (55.6%), individuals reporting severe stressful life events (25.4%), those with other substance use (42.6%), and participants with depressive symptoms (PHQ-9 ≥10; 40.8%).

### Associated factors of problematic khat use

In the bivariable analysis, female participants had lower odds of problematic khat use than males (COR = 0.21, 95% CI: 0.07–0.58). Daily or almost daily khat users had 9.70 times higher odds than occasional users (COR = 9.70, 95% CI: 5.22–18.02). After adjusting for potential confounders, daily or almost daily khat users had 8.05 times higher adjusted odds compared with occasional users (AOR = 8.05, 95% CI: 3.75– 17.24). Participants with stressful life events had 3.48 times higher adjusted odds (AOR = 3.48, 95% CI: 1.56–7.76), and those with depressive symptoms had 3.70 times higher adjusted odds of problematic khat use (AOR = 3.70, 95% CI: 2.00–6.75). The associations observed for sex, other substance use, employment status, and other substance use in the crude analysis were no longer statistically significant in the adjusted model.

## Discussion

This community-based cross-sectional study provides important evidence on the burden of problematic khat use among adults in Ethiopia. Using the Problematic Khat Use Screening Test (PKUST-17), a locally developed and validated instrument, we found that 19.6% of users had problematic khat use. This is the first time we have examined potential determinants for the transition from khat use to problematic khat use. After adjustment, daily or almost daily khat use, severe stressful life events, and depressive symptoms were independently associated with problematic khat use. Together with our previous work(Mihretu et al., 2022), these findings add to the emerging evidence that problematic khat use exists and represents an important public health concern in Ethiopia.

Khat use is substantial in Ethiopia, with national estimates rising from approximately 16% to 23% in recent surveys(Ethiopian Public Health Institute (EPHI), 2025; Teklie et al., 2018). However, the proportion of users who develop problematic use has received little attention. Our estimate of 19.6% is close to the 14% reported in the Ethiopian Somali Region, with the variation reflecting differences in culture, availability, and consumption patterns.

International evidence also supports the high prevalence of khat dependence. Studies from Yemen, Saudi Arabia, and European migrant communities have reported psychological dependence among 30–50% of khat users (El-Setouhy et al., 2016b; Griffiths et al., 1997; Kassim et al., 2013). Although prevalence varies across settings, the consistent finding is that a substantial proportion of khat users develop dependence symptoms and functional impairment. In our study, problematic use was particularly common among men, daily users, and those experiencing psychosocial adversity.

PKUST-17 adds to the growing evidence for culturally appropriate assessment of problematic khat use. Previous validation studies have demonstrated that it measures a distinct construct associated with dependence symptoms, stressful life events, depression, and other substance use (Mihretu et al., 2022a, 2025). Its use in community settings supports its relevance for both research and clinical screening.

The strong association between daily khat use and problematic use is biologically and socially plausible. Cathinone, the main psychoactive component of khat, stimulates dopaminergic reward pathways, while repeated exposure may contribute to tolerance, psychological dependence, and withdrawal symptoms (Kalix, 1992). At the same time, the widespread social acceptance and availability of khat may facilitate frequent consumption. Qualitative studies from Ethiopia similarly show that users distinguish problematic use by loss of control, continued use despite harm, and using khat to avoid withdrawal (Mihretu et al., 2017).

We also found a strong association between problematic khat use and depressive symptoms, but the direction of this relationship remains uncertain and is likely bidirectional (Mihretu et al., 2025; Warfa et al., 2026). Individuals with depression may use khat as a coping strategy, whereas prolonged and problematic use may worsen sleep and emotional regulation, and then overall psychological well-being. Similarly, severe stressful life events may increase vulnerability to harmful patterns of use through maladaptive coping mechanisms.

These findings have important clinical and public health implications. Early identification of daily khat users may help prevent progression to problematic use. Integrating PKUST-17 into primary healthcare, mental health, and substance use services could enable routine screening, brief psychological interventions, motivational interviewing, and treatment of co-occurring mental health conditions.

The study’s main strength is its use of a locally developed, psychometrically validated screening tool with strong cultural relevance. However, PKUST-17 identifies probable problematic khat use rather than a formal DSM-5 diagnosis. Self-reported substance use may also be affected by recall and social desirability bias. The relatively small sample size in some cells resulted in wide confidence intervals and reduced estimate precision.

## Conclusion

Problematic khat use affects approximately one in five khat users in Ethiopia and is strongly associated with daily khat use, stressful life events, and depression. Future longitudinal studies are needed to clarify causal pathways and to evaluate culturally appropriate prevention and treatment interventions.

## Data Availability

The manuscript includes all the relevant data.

---

**Table 1:**
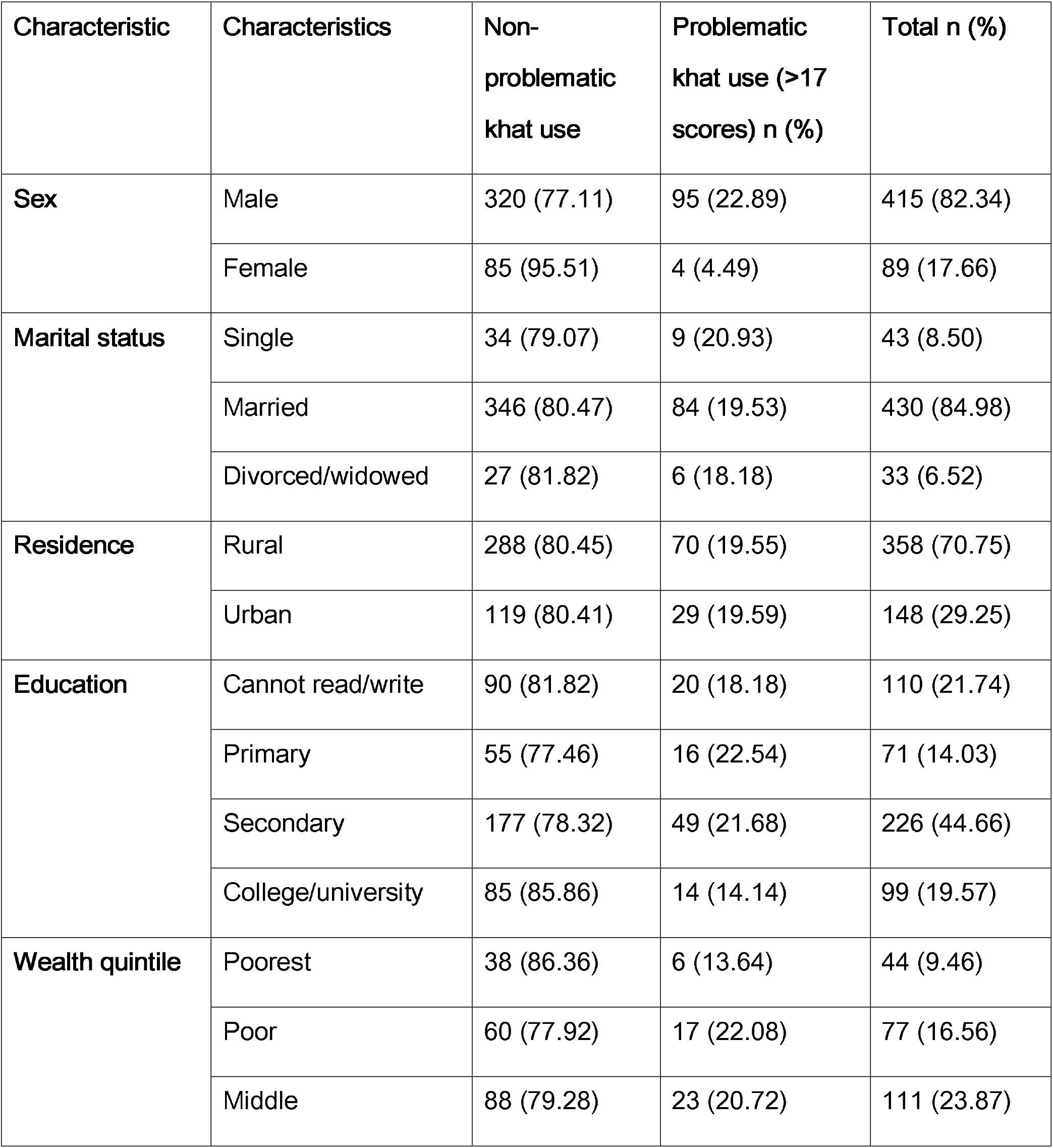
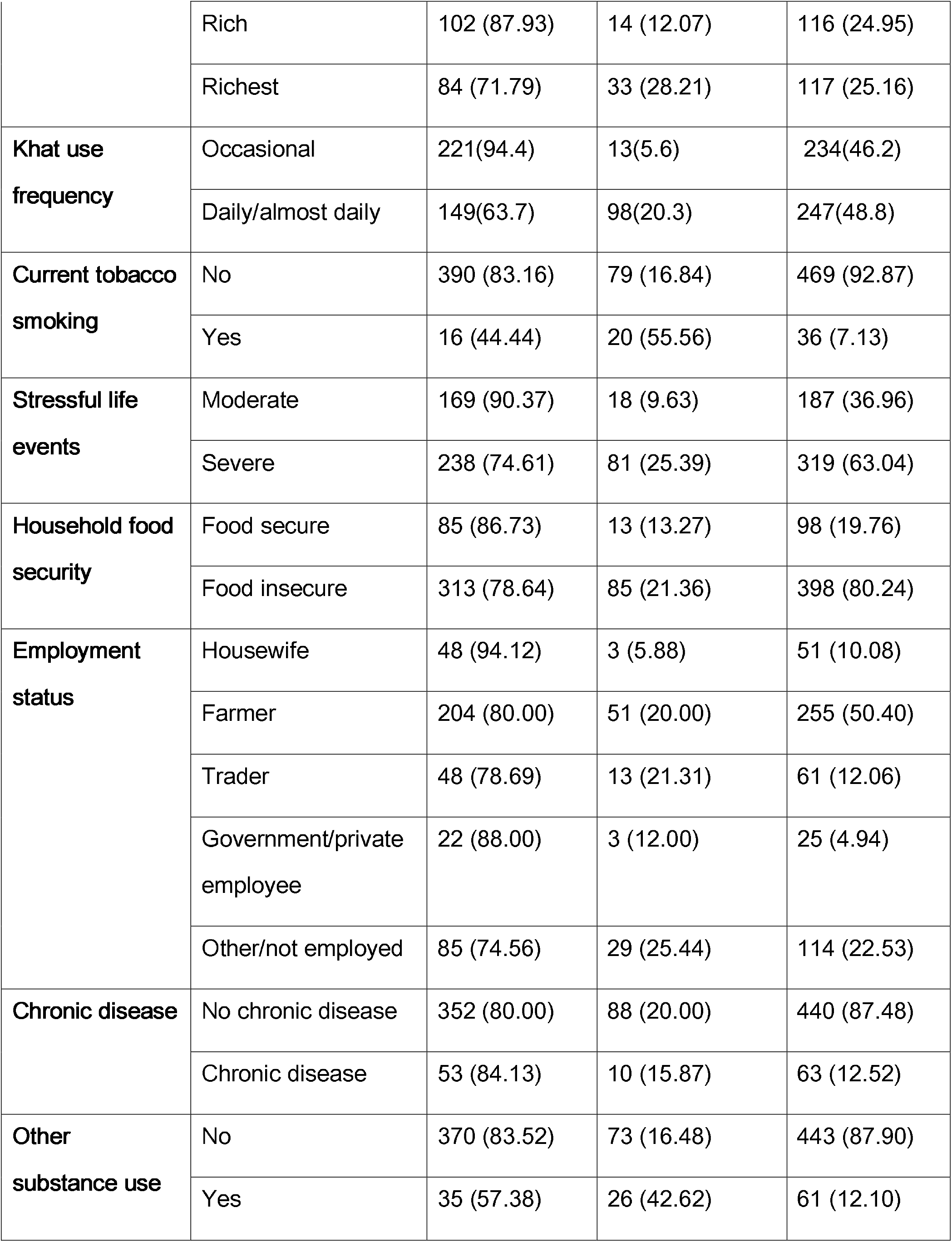
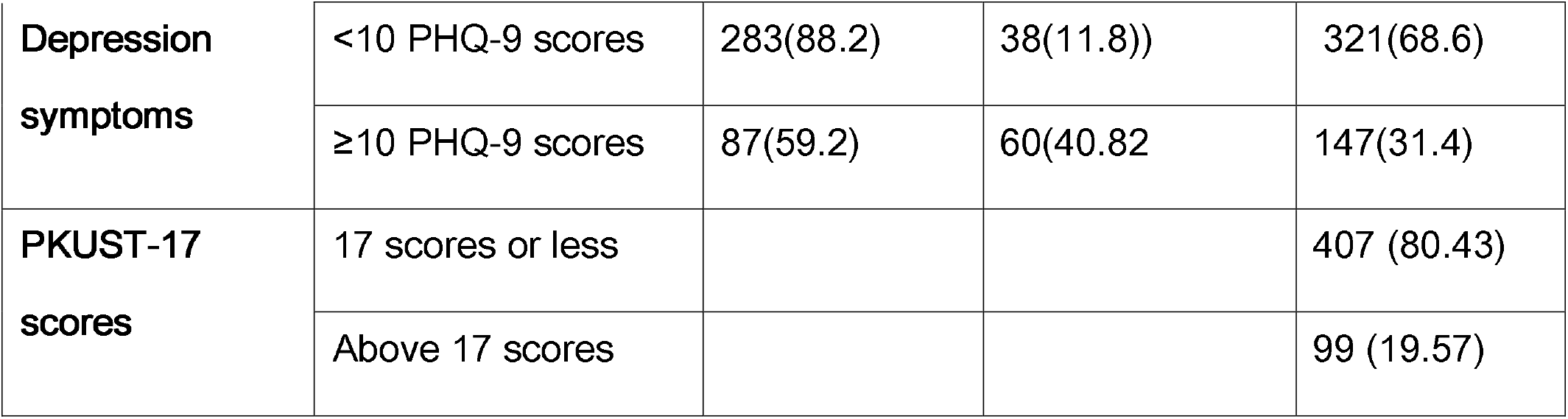
Sociodemographic and behavioural characteristics by PKUST-17 status (n=504)

| Characteristic | Characteristics | Non-problematic khat use | Problematic khat use (>17 scores) n (%) | Total n (%) |
| --- | --- | --- | --- | --- |
| **Sex** | Male | 320 (77.11) | 95 (22.89) | 415 (82.34) |
|  | Female | 85 (95.51) | 4 (4.49) | 89 (17.66) |
| **Marital status** | Single | 34 (79.07) | 9 (20.93) | 43 (8.50) |
|  | Married | 346 (80.47) | 84 (19.53) | 430 (84.98) |
|  | Divorced/widowed | 27 (81.82) | 6 (18.18) | 33 (6.52) |
| **Residence** | Rural | 288 (80.45) | 70 (19.55) | 358 (70.75) |
|  | Urban | 119 (80.41) | 29 (19.59) | 148 (29.25) |
| **Education** | Cannot read/write | 90 (81.82) | 20 (18.18) | 110 (21.74) |
|  | Primary | 55 (77.46) | 16 (22.54) | 71 (14.03) |
|  | Secondary | 177 (78.32) | 49 (21.68) | 226 (44.66) |
|  | College/university | 85 (85.86) | 14 (14.14) | 99 (19.57) |
| **Wealth quintile** | Poorest | 38 (86.36) | 6 (13.64) | 44 (9.46) |
|  | Poor | 60 (77.92) | 17 (22.08) | 77 (16.56) |
|  | Middle | 88 (79.28) | 23 (20.72) | 111 (23.87) |
|  | Rich | 102 (87.93) | 14 (12.07) | 116 (24.95) |
|  | Richest | 84 (71.79) | 33 (28.21) | 117 (25.16) |
| **Khat use frequency** | Occasional | 221(94.4) | 13(5.6) | 234(46.2) |
|  | Daily/almost daily | 149(63.7) | 98(20.3) | 247(48.8) |
| **Current tobacco smoking** | No | 390 (83.16) | 79 (16.84) | 469 (92.87) |
|  | Yes | 16 (44.44) | 20 (55.56) | 36 (7.13) |
| **Stressful life events** | Moderate | 169 (90.37) | 18 (9.63) | 187 (36.96) |
|  | Severe | 238 (74.61) | 81 (25.39) | 319 (63.04) |
| **Household food security** | Food secure | 85 (86.73) | 13 (13.27) | 98 (19.76) |
|  | Food insecure | 313 (78.64) | 85 (21.36) | 398 (80.24) |
| **Employment status** | Housewife | 48 (94.12) | 3 (5.88) | 51 (10.08) |
|  | Farmer | 204 (80.00) | 51 (20.00) | 255 (50.40) |
|  | Trader | 48 (78.69) | 13 (21.31) | 61 (12.06) |
|  | Government/private employee | 22 (88.00) | 3 (12.00) | 25 (4.94) |
|  | Other/not employed | 85 (74.56) | 29 (25.44) | 114 (22.53) |
| **Chronic disease** | No chronic disease | 352 (80.00) | 88 (20.00) | 440 (87.48) |
|  | Chronic disease | 53 (84.13) | 10 (15.87) | 63 (12.52) |
| **Other substance use** | No | 370 (83.52) | 73 (16.48) | 443 (87.90) |
|  | Yes | 35 (57.38) | 26 (42.62) | 61 (12.10) |
| **Depression symptoms** | <10 PHQ-9 scores | 283(88.2) | 38(11.8)) | 321(68.6) |
|  | ≥10 PHQ-9 scores | 87(59.2) | 60(40.82 | 147(31.4) |
| **PKUST-17 scores** | 17 scores or less |  |  | 407 (80.43) |
|  | Above 17 scores |  |  | 99 (19.57) |

**Table 2:**
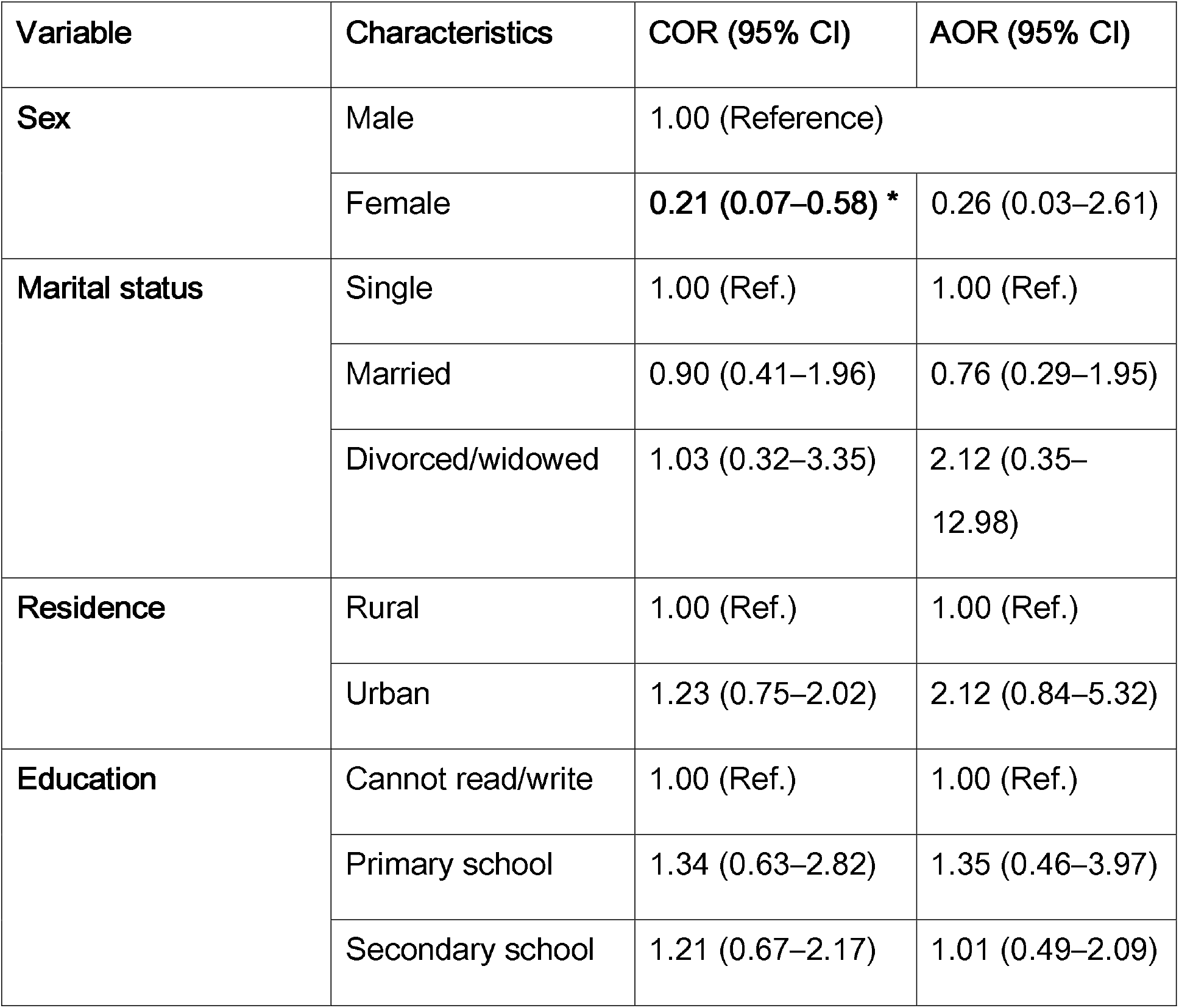
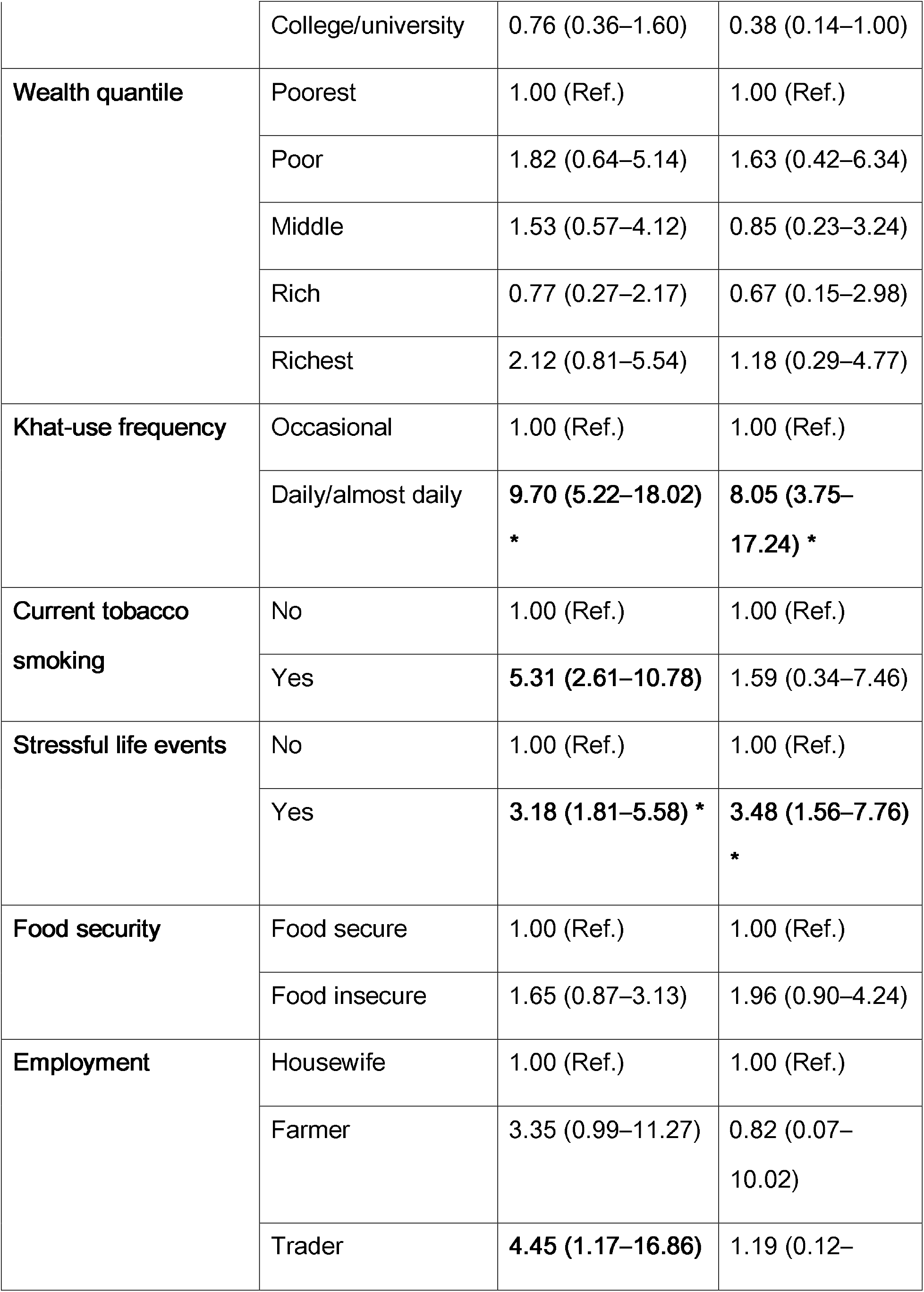
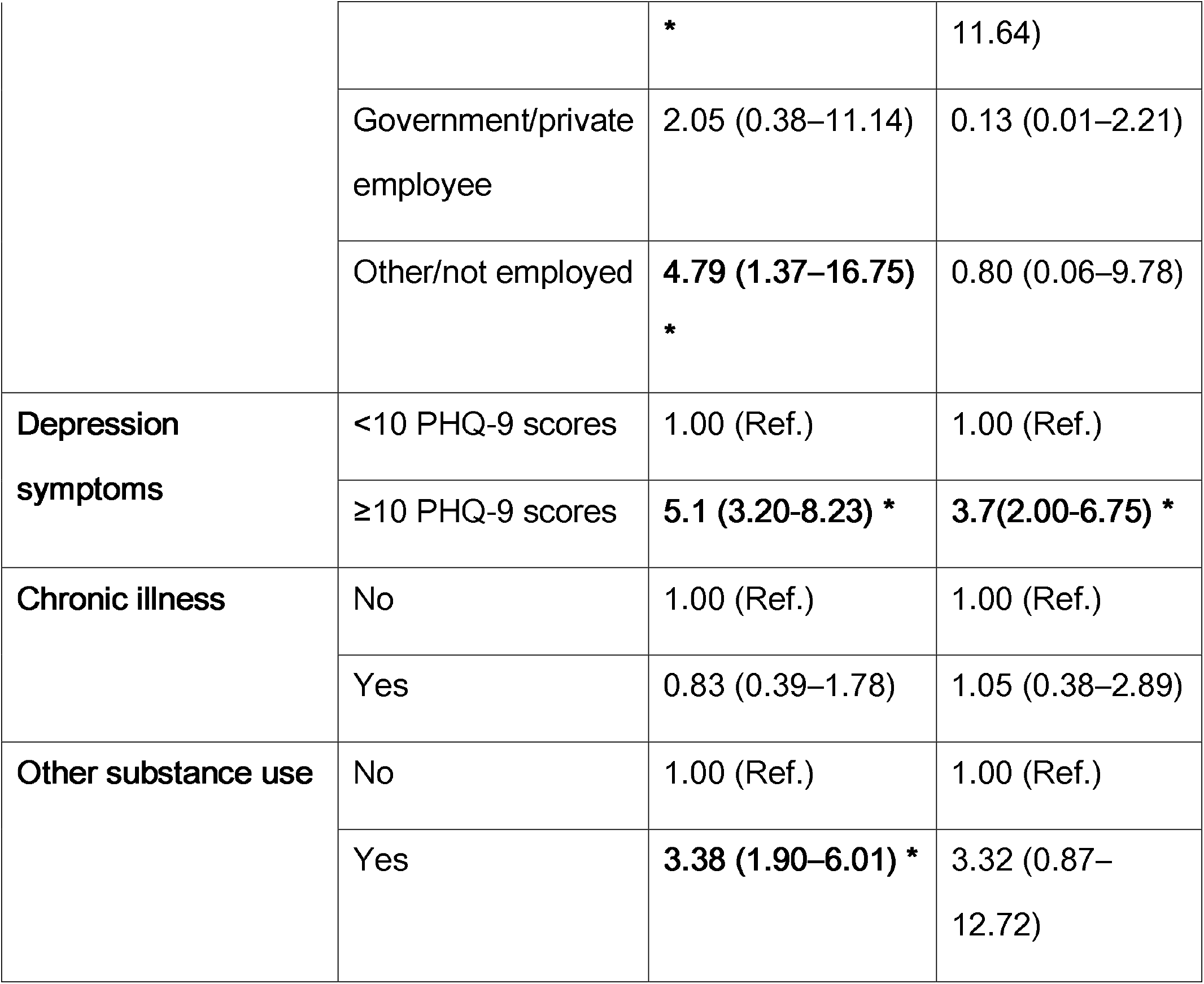
Crude and adjusted odds ratios for factors associated with problematic khat use among khat users (n=504)

## Notes

### Competing Interest Statement

The authors have declared no competing interest.

### Author Declarations

The study was approved by the Institutional Review Board of the College of Health Sciences, Addis Ababa University (Reference: 023/24/Psy).

